# Informal caregiving following stroke: a qualitative exploration of carer self-identification, care-related language and support experiences

**DOI:** 10.1101/2024.10.07.24315049

**Authors:** Bethany J. Harcourt, Richard J. Brown, Audrey Bowen

## Abstract

**Background and objectives:** Following stroke, people often receive informal care from friends and family. Some carers adopt the role quickly whereas others find it more difficult to see themselves as a ‘carer’ and do not access relevant support. This project sought to understand the process of how and when informal carers start to see themselves as such, as well as their preferred terminology and experiences of support in this role.

**Methods:** Qualitative semi-structured interviews with eighteen UK adults who had provided care or support to a friend or family member after a stroke. Data were analysed thematically using a template analysis approach. PCPI collaboration, through a formed carer advisory group, enhanced the study methodology.

**Results:** Five main themes were developed: *1) Adopting and adjusting to the care role, 2) Accessibility of support, 3) Perceptions of support, 4) Acceptability of care-related language and terminology, 5) Function of care-related language and terminology.* Facilitators and barriers to participants self-identifying with the care role were identified. Self-identification was found to commonly occur at four key points along the stroke trajectory. Two main contrasting opinions around the acceptability of care-related terminology were shared. Accessibility of support services and suggestions for future support provision were discussed.

**Conclusion:** Individuals identify as ‘carers’ at different points and to different extents throughout the stroke trajectory. Various factors influence this process. These findings have implications for the provision of more inclusive and accessible support for informal caregivers of stroke survivors.

**Strengths and limitations of this study:**

- This study featured heterogeneity in the participant sample in terms of age, relationship to the stroke survivor and duration of care experience.
- The use of collaboration with a PCPI group strengthened the study, including with accessibility and appropriateness of the study to participants.
- The retrospective approach to the study enabled participants to reflect of their caregiving experience, their insights, and perspectives they developed through this journey.
- This study is limited by its minimal diversity in terms of participant ethnicity.

## 1. Introduction

Stroke is a sudden onset neurological condition that can have significant impacts on physical, cognitive and sensory functioning [1]. Central to stroke survivors’ rehabilitation and recovery is often the provision of care and support from individuals close to them [2].

The Stroke Association defines a carer as “someone who provides unpaid help and support to family and friends” [3]. The term ‘informal carers’ is often used to distinguish this population from carers in paid employment. The informal caregiving experience can differ between individuals, including both positive and challenging aspects [4]. After undertaking this role, carers have significant needs and require appropriate support [5–7]. The Care Act establishes the role of local authorities to assess carers’ needs when they request this and plan any required support [8].

Adopting the ‘carer’ role can be a significant change to one’s identity. Emotional and psychological difficulties can occur following this, such as increased anxiety and stress, and an increased sense of responsibility [9]. The psychological adjustment to becoming a carer can include restructuring life, responsibilities, and family dynamics to accommodate this role [10]. Informal carers do not always identify as ‘carers’ or use this language to describe themselves [11]. Within caregiving across other neurological and long-term health conditions, individuals commonly express a preference for being affiliated and recognised with their relationship role (e.g. ‘partner’), rather than being described as a ‘carer’ [12–13]. Nevertheless, even individuals who do not initially identify as a ‘carer’ often reach a point of recognition and acceptance of their care-providing role, even if they do not use the term ‘carer’ [14–15].

Limited current research explores the process of self-identification for informal carers of stroke survivors, and how and when they come to see themselves as such. This has been explored to an extent in other populations and neurological conditions. A staged transitional caregiving framework was developed with new carers of people with dementia and chronic health conditions to guide carer-focused support provision [16]. An extension of this framework, caregiver identity theory, emphasises the dynamic nature of caregiving, and how carers’ identity can evolve throughout this experience [16].

Relating to stroke, the ‘Timing it Right’ framework identifies five core stages of caregiving for informal carers, from stroke onset to the point of adaptation to the role [17]. This describes some elements of post-stroke life that informal carers may adjust to, such as changes to family dynamics and routines and learning and implementation of care-related skills. However, it does not specifically focus on the carer’s changing sense of identity. A limited focus on the self-identification process to this new identity is common to other theories in this population [9,18–19].

Investigating the process of carer self-identification is of core interest and relevance to the profession of clinical psychology. Clinical psychologists play a key role in supporting stroke survivors, their family and informal carers with emotional and psychological difficulties following stroke [20]. This includes supporting carers’ psychological adjustment to their role [21]. Increasing the understanding of carers’ psychological adjustment to a changing sense of identity and identifying factors influencing this process could aid this support provision. Understanding *when* this self-identification happens could inform the profession with regards to the timing of offering support.

Furthermore, clinical psychologists are positioned in stroke services to provide training, clinical supervision, advice and support regarding psychological impacts of stroke to multidisciplinary team members [22]. Better information relating to the informal caregiving experience and associated psychological adjustment processes may strengthen clinical psychologists’ ability to support the wider multidisciplinary team in supporting and meeting the needs of this population.

Clinical psychologists are often involved in service and intervention development and evaluation [23]. Studies investigating the effectiveness of person-centred interventions for informal carers of stroke survivors identified challenges in implementing support for this population, including carers having difficulty recognising their own needs, and often not seeing themselves as ‘carers’ [11]. This may contribute to difficulties accessing and engaging with healthcare professionals for support [11, 24]. Further research on carer self-identification is required to understand the extent to which this impacts support provision accessibility, with a view to shaping the future support offered to this population.

Furthermore, exploring this topic with the aim of improving the support provision for informal carers of stroke survivors aligns with agreed research objectives of the James Lind Alliance (JLA) Stroke Priority Setting Partnership [25]. The JLA enable clinicians, patients and carers to work collaboratively to identify and prioritise areas of research and evidence uncertainty in healthcare [26]. Improvements to the carer experience is identified as a top 10 priority for rehabilitation and long-term care in stroke research to guide intervention and support provision [25]. Therefore, exploring this area has the potential to contribute towards this evidence base.

### Study objectives

This study aims to answer three inter-related questions regarding self-identification in informal caregivers of stroke survivors: (1) How and when does someone see themselves as a ‘carer’ or self-identify as a person providing care to someone after stroke? (2) What care-related language have participants experienced and what are their preferences around this? (3) What has the experience of support been for participants?

## 2. Method

### 2.1. Design

Qualitative semi-structured interviews were conducted with consenting informal carers of stroke survivors. Data were analysed using a template variant of thematic analysis [27]. Ethics approval was obtained from the University of Manchester’s Research Ethics Committee. (Ref: 2023-15237-27180).

### 2.2. PCPI Involvement

Patient, carer and public involvement (PCPI) [28] was important to the project. Liaison with Ann Bamford, Lay Lead for PCPI in the Geoffrey Jefferson Brain Research Centre, helped develop an appropriate and inclusive recruitment strategy. A carer advisory group was formed by the lead researcher of individuals with experience providing informal care to a stroke survivor. This group was engaged in a collaborative capacity [29] in project planning (i.e. strategies for recruiting a diverse sample, accessibility and appropriateness of study materials), data analysis (i.e. sharing of fully anonymised quotes and data extracts to aid the research team’s interpretation of the data) and dissemination plans (i.e. accessibility of information formats).

### 2.3. Participant eligibility and recruitment

Participants had to be UK residents aged ≥18 years and to have provided informal care to an individual who had experienced a stroke at least one year ago. The stroke being at least one year ago allowed sufficient time for participants to reflect on their experiences whilst minimising any risk of distress discussing these. The recruitment strategy used several channels and sampling methods [30] to recruit a diverse and varied participant sample, in terms of age, ethnicity and gender, and thereby reduce the risk of under-represented populations in research being omitted from the study [31]. Local and national third sector organisations were contacted via email, telephone and in-person. These included stroke charities and organisations, social groups, wellbeing spaces, carer groups and forums, faith groups, and groups and organisations representing and supporting ethic minority populations. Organisations such as shops, supermarkets, gyms, and leisure centres across diverse regions were also contacted and visited to advertise the study. The lead researcher offered to complete talks about the project at events or activities being run by these organisations. The research team also shared details of the study through their professional networks and via professional social media accounts.

### 2.4. Materials and procedure

Participants expressed interest via email or telephone contact with the researcher. Information sheets were provided via email or post, depending on participants’ preferences. Telephone or email contact was made to confirm interest and eligibility and schedule interviews. After understanding the principle of limited confidentiality and providing written or audio-recorded consent, participants provided demographic information, including age, gender identity, ethnicity and stroke-related information.

Qualitative data were then collected using semi-structured interviews with open-ended questions, allowing for an in-depth exploration of participants’ views and experiences [32]. Preferred terminology (i.e. carer, caregiver, family member) was discussed at the start of interviews to promote inclusivity and consideration of participants’ preferences, and interviewer-participant rapport. The interview topic guide was developed in liaison with the study’s carer advisory group. Areas the interview focussed on included participants’ experience and preferences around care-related language, the process and timing of identifying with the caring role and facilitators and barriers to this, and their experience and accessibility of support.

Interviews were audio-recorded and transcribed by the lead researcher. All identifiable information was removed from the transcripts, which were uploaded to NVivo 12 software [33] for analysis. On interview completion, participants were thanked and provided with a debrief sheet, including signposting to relevant support services.

### 2.5. Data Analysis

Qualitative template analysis [36] was chosen as an appropriate data analysis method as it encompassed both deductive and inductive approaches. A contextual constructivist position was adopted, which assumes there are always multiple interpretations to be made of any phenomena, which can depend on the researcher’s position and social context in which the research is conducted [35]. Adopting this position allowed *a priori* themes to be posed more tentatively than in other epistemological positions yet allowed the data to drive the analysis without being overly constrained by these existing constructs and themes. A priori themes were constructed from established theoretical frameworks on stages of caregiving [17] and reviews of existing literature. These were then modified and added to inductively, as led by the data, providing the richest quality of data into perspectives of the target population.

The researcher immersed themself in the data by thoroughly re-reading and familiarising themselves with each transcript. In line with template analysis approaches [36], initial line-by-line coding was completed on a subset of six transcripts (i.e., one third of the total). Data segments that were deemed relevant to research questions were identified. If these segments were embodied within an *a priori* theme, that code was assigned. Where relevant segments were not encapsulated by an a priori theme, new codes were developed, and existing codes modified flexibly.

Discussions within the research team regarding initial transcripts early in the analysis process promoted rigour. After coding the first six transcripts, the initial coding template was discussed reflectively with the research team. The coding template was comprised of higher order codes that were present across multiple transcripts and represented broader themes across the data set, in addition to other levels of coding, applied to more specific or less prevalent themes. As the remaining transcripts were coded and analysed, the template was further developed in an iterative process. The final coding template had three coding levels. This was then applied to the full data set.

### 2.6. Rigour

The first two transcribed interviews were reviewed within the research team, to reflect on the interview schedule effectiveness in addressing the research questions. The lead reviewer (BH) coded the first six interviews and developed the initial coding template. The coded data and initial coding template were shared with another author (AB) to clarify the template before coding the remainder. The research team (BH, RB, AB) reviewed further coding template iterations and explored, through reflective discussions, the relevance and development of *a priori* themes as coding procee_ded_. This allowed the narrative to be developed from the data, and for the coding template refinement and modification to be an iterative process. These discussions enabled exploration of alternative ways of making sense of the data, and how this could be reflected through the coding template organisation. PCPI input promoted study rigour through an independent scrutiny of data interpretation, a recommended quality check within the template analysis approach [37].

### 2.7. Reflexivity

During the research process the lead researcher held an awareness of, and reflected on, their personal and professional identity, roles, and assumptions. The lead researcher was mindful of their personal experience and professional interest in informal caregiving for other neurological conditions. Through keeping a reflective log and supervisory research team discussions, they reflected on how this component of their identity was beneficial to building rapport with participants, whilst needing to be held in mind throughout the research process to minimise potential influences on interview questioning and data interpretation.

## 3. Results

### 3.1. Participant Sample Characteristics

Eighteen individuals were recruited to participate. Interviews lasted approximately 54-89 minutes (Mean length= 69 minutes) and were conducted via telephone (n=2) or Zoom (n=16). Recruitment concluded after 18 interviews because the existing interviews provided sufficient information depth and repetition to address the research questions [38].

Participant age ranged widely from 19-92 years (mean = 58 years). Most participants (n= 16, 89%) identified as White British, one participant identified as mixed ethnicity, White British and Black African, and another identified as Asian. The nature of the relationship to the stroke survivor, education and employment history varied across participants, most of whom were women (Table 1).

**Table 1:** Overview of demographic characteristics of participants.

| Demographic Criteria | Participant Information<br>n (%) |
| --- | --- |
| **Gender Identity** |  |
| Female | 13 (72) |
| Male | 5 (28) |
| **Age, mean** | 57.8 |
| **Disability Status** |  |
| Disabled | 1 (6) |
| Non-disabled | 17 (94) |
| **Employment Status** |  |
| Full-time employment | 4 (22) |
| Part-time employment | 3 (17) |
| Other- including full time carer and mother | 3 (17) |
| Retired | 7 (39) |
| Student | 1 (6) |
| **Highest level of education** |  |
| Examinations at 16 | 3 (17) |
| College/Sixth form or equivalent | 6 (33) |
| University Undergraduate degree | 8 (44) |
| University Masters degree | 1 (6) |
| **Relationship with stroke survivor** |  |
| Spouse/partner | 10 (56) |
| Brother/Sister | 1 (6) |
| Son/daughter | 5 (28) |
| Grandson/granddaughter | 1 (6) |
| Friend | 1 (6) |
| **Lives relative to stroke survivor** |  |
| In the same household | 8 (44) |
| In different household | 10 (56) |
| **Months post stroke (at time of interview)** |  |
| Mean (min.- max. range) | 103 (12-348) |
| **Previous experience of providing care prior to stroke** |  |
| Yes | 8 (44) |
| No | 10 (56) |
| **Number of people currently providing care for** |  |
| Mean (min.- max. range) | 1 (0-3) |

### 3.2. Qualitative findings

Results were organised into five main themes: *1) Adopting and adjusting to the care role, 2) Accessibility of support, 3) Perceptions of support, 4) Acceptability of care-related language and terminology, 5) Function of care-related language and terminology.* All participants provided data relating to each main theme, yet experiences and perspectives varied.

Theme 1 was comprised of two further levels of subthemes, and themes 2, 3 and 5 each had one subtheme level. A thematic diagram is displayed in Figure 1. Template analysis includes more coding levels than other qualitative thematic analysis methods [24]. Therefore, to meaningfully and coherently present findings, themes will be drawn on to answer this study’s three research questions via a deductive approach. This results presentation is consistent with other template analysis approach qualitative studies [39–40].

**Figure 1:**
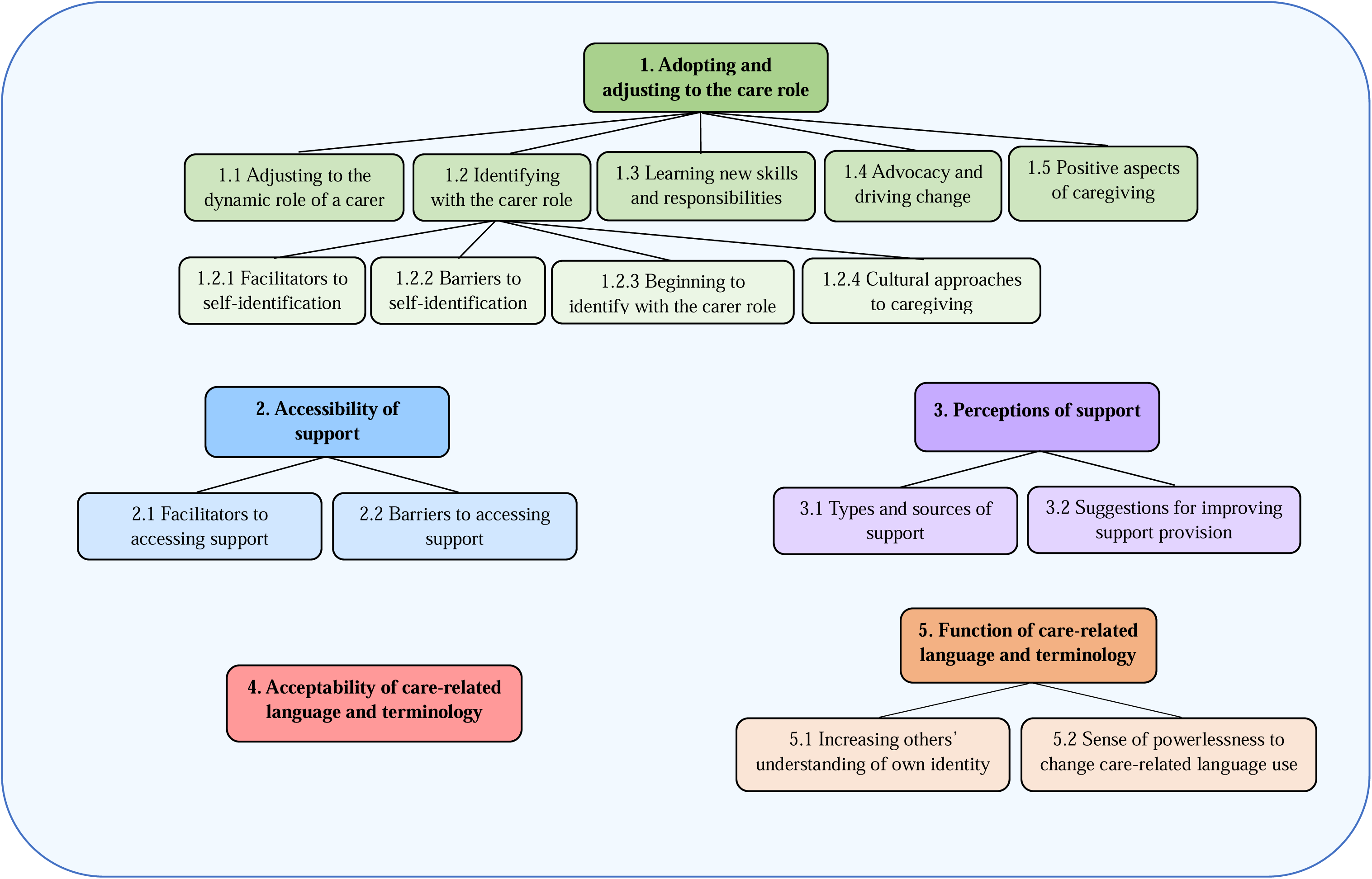
Visual depiction of themes and subthemes.

#### ‘How’ and ‘when’ do people self-identify as providing care for stroke survivors?

The generated understanding addressing this research question is drawn from *Adopting and adjusting to the care role* (Theme 1) and *Identifying with the carer role* (Subtheme 1.2). Subthemes 1.2.1-1.2.3 relate to the components of *how* and *when* this self-identification process occurs.

In exploring *how* participants came to self-identify with their care role, they all discussed *Facilitators to self-identification* (Subtheme 1.2.1). A common factor participants identified as facilitating this process was the stroke impact. Participants expressed that through involvement in rehabilitation processes, physically seeing or being made aware of (e.g. through liaison with healthcare professionals) the stroke impact and their loved one’s impairments moved them towards a position of acknowledging they may need to undertake a care-related role.

> *“…just from me going in and seeing what he could and couldn’t do and seeing things like he hadn’t been for a shower maybe, or got changed. So I would be like “Right, come on let’s do this together”. So I think in terms of first identifying me in a caring role for him was a bit of both really, both the professionals at the hospital and myself, just from my own observations really.” (Participant 10)*

Another facilitator to self-identification was connecting with others in a similar situation. Participants able to access this peer support discussed how it facilitated their recognition of their changed role. Participants who lacked access to peer support felt it could have helped them self-identify with and adjust to their new care role.

> *“I was with my sister whose husband is ill, and she is an unofficial carer for him as well. And we both sort of looked at each other and said “Oh goodness! We’re both carers aren’t we!”. So that moment of realisation didn’t happen when I was in a surgery or a hospital environment or whatever, it was just a realisation that we were both doing all the things that a carer would do.” (Participant 3)*

Participants reflected on how recognition from other people of the supportive role they had taken on helped them to start to see themselves as carers. Participants expressed that in hindsight, they often struggled to identify with the care role by themselves and in the moment, yet the recognition and validation from others helped them to recognise and identify more with their changed role. This included friends, family members, and third sector support organisations.

*“I think when my friends recognised what I was doing for my dad and told me that they thought I was his carer, it affected me immediately, you know, straight away. It made me recognise that yeah, that was what I was and that was what I was doing for him.” (Participant 14)*

Changes to family system and relationship dynamics following stroke were also identified as a factor that helped participants start to identify more with the care-providing role.

> *“He is basically the whole rock of the family. So the minute he went down, it was like the queen. The minute he went down, everyone around him just went into that short sharp shock reality of “right, we’ve got to look after granddad”(Participant 7)*

This included when changes occurred within the relationship between the participant and the stroke survivor, such as the nature and type of the relationship.

> *“I would always see myself and identify more as a carer now than as a partner. Because there’s not much intimacy or anything like that anymore, so that makes me feel more like a carer and identify more as that.” (Participant 18)*

Another facilitator to self-identification was participants feeling that they were the only person able to take on the role. This factor was particularly salient for those who identified as having limited family support. These participants described an increased sense of responsibility and felt this contributed to them identifying with the carer role sooner than they may have otherwise.

> *“I identified myself as a carer straightaway, because I think I knew that I had to be the one to step up because there was no one else. I think that’s the only reason that I accepted it at the beginning. I knew he needed this help and support, and there was nobody to provide that except me.” (Participant 18)*

The concepts of *learning new skills and responsibilities* (Subtheme 1.3), and *advocacy and driving change* (Subtheme 1.4) were discussed by all participants as distinct experiences within informal caregiving yet were also identified as contributing to their developing self-identity. This included learning to adapt communication, especially if their loved one had developed speech difficulties or aphasia.

> *“I became an interpreter. I’ve been there when physios have been there and they’ll be asking questions…and my dad would be waffling away and they’d be looking at me confused, and so I would have to interpret and say, “Oh he’s saying this, this and this”. It was almost like interpreting another language really that only I understood, because you’ve just got that level of understanding between one another” (Participant 14)*

Motivating the stroke survivor was an important skill participants described having developed, to support their wellbeing and rehabilitation progress. Participants felt this new responsibility was adopted as part of their care role, and was outside the scope of their relationship role. It was therefore identified as a facilitator to carer self-identification.

> *“It did sort of give you a purpose as a carer…It was the fact that you were part of a system to try to restore the person that you loved back to some semblance of normality.” (Participant 6)*

> *“So, at times when he gets frustrated and thinks he isn’t progressing or hasn’t progressed, I think right, well we need to stop and reflect here and just really see what has happened and now where you’re at.” (Participant 17)*

Many participants described adopting a role of *advocacy and driving change* for the stroke survivor. Some participants spoke about assuming the advocacy role willingly and readily to ensure their loved one’s needs were met and service provision was accessible.

Participants who discussed this subtheme expressed how the need for them to engage with advocacy strengthened their affinity to the care role identity.

> *“I used to be quite laid back and then I found that if you didn’t shout for things, then you didn’t get them. I was shouting for things for mum, not for me…I suppose that I signed up for the carer job and so it’s just what I did” (Participant 4)*

Some participants discussed *positive aspects of caregiving* (Subtheme 1.5), such as experiencing a sense of pride or achievement in supporting their loved one. They reflected on how this helped them feel a stronger connection to their care role identity.

> *“Well I think it might come down to pride to be honest. I was quite proud of what I did and hearing those comments from my friends helped me feel proud. There are positives to providing care for somebody.” (Participant 12)*

> *“It’s the extended role of actually having to support somebody…I feel so much reward from it, and I don’t even do it full time. Like seeing my dad when he first, well when there was milestones he had, like when he got home, and on the first anniversary of his stroke I felt so much like, ‘Oh, my God! Like he’s made it! We never thought this was going to happen’, and although it has, like the awful psychological effects, you also get such benefits from it in terms of you feel so proud of what you’ve done and you also feel so proud of what your father has accomplished.”(Participant 13)*

> *“I do think you can get a kick out of it… it can build you up as a person and give you strength of* character*.” (Participant 12)*

Participants often spoke about relationships strengthening between themselves, the stroke survivors, family members and making new friends along their journey because of the shared experience.

> *“Going through that brought us closer together.” (Participant 5)*

In exploring *how* participants reached the point of self-identifying with the care role, it was important to explore any *barriers to self-identification* (Subtheme 1.2.2), that made it more difficult to see themselves as a ‘carer’ or as someone providing care. Due to the sudden increase in responsibility and the taking on of a new role, participants often described a feeling of life feeling too busy or chaotic for them to have the time to stop, think or reflect on their experiences. Many participants felt this contributed to a delayed recognition of their changed role and level of care and support they were providing.

> *“But at the time I don’t think people often think about it, as you just get on with it and you’re just doing it and don’t really have time to stop and think about yourself or your role.”(Participant 16)*

Reflecting on the experience of assuming a care role, participants highlighted that they often neglected or did not prioritise their own needs during the initial stages. They discussed how this included not realising the impact of the stroke on themselves, their resultant needs, and how they saw themselves. Therefore, participants thought this further contributed to the delay in recognising their care role and the scope of this.

> *“Now looking back I think it’s kind of obvious that I was a carer and I think I probably should have realised that. But at the time it just didn’t occur to me because I was so busy. You just don’t have time to actually stop and think or even process what is happening to you. So I just didn’t realise I was a carer for a while. At the time it was just too manic.”(Participant 16)*

Participants discussed *adjusting to the dynamic role of a carer* (Subtheme 1.1) as a broad concept, in addition to as a barrier to self-identification. They reflected on how they were not adjusting to a static role, but one that changed and transitioned through the stroke trajectory in response to stroke survivors’ changing needs. This often contributed to a sense of uncertainty and unpredictability that participants felt took time to process and adjust to, meaning it could take time for them to recognise the scope of their care role and identify with this.

> *“The problem with sailing a yacht is that you need to know where the wind direction is coming from to set your sails and, well it’s a bit like that with caring, sometimes the wind is coming from all directions at once and you’re just left wondering what move to make next.” (Participant 12)*

The language and terminology used around providing care constituted a significant barrier for many participants in both their adjustment to the role and to them reaching a point of self-identification. One element to this was participants’ perception of the carer role. Participants often showed a reduced understanding of the difference between formal (paid) and informal (e.g. family, friend) carers, and often had a specific idea of what a ‘carer’ was, that may feel unrelatable to them.

> *“I’m not really like helping him get dressed, I’m not feeding him, I’m not doing all this and that. So, it is not something I necessarily would call myself.” (Participant 13)*

This included them feeling that adopting the role of a “carer” meant a permanent change to their identity, which some participants didn’t feel reflected their changing needs and experiences throughout the stroke trajectory.

> *“I also think that the word carer is often seen as a permanent thing rather than a moveable thing. People might think that you are now a carer, and now that is your role. So they don’t see that it’s a moveable thing, or that people might stop becoming carers or identify less as a carer as that person gets better.” (Participant 16)*

When they were supported with education and information around the diversity and vast scope of a care role, this helped participants understand and identify more with this.

> *“To me a carer was somebody who puts the uniform on, goes off to work and does the caring, then comes back and takes the uniform off kind of thing. But it’s not. So I had to come to terms with it in my own mind and reassess actually, “What is a carer?””(Participant 11)*

Furthermore, some participants spoke about enduring lack of understanding and stigma at a societal level of the informal caring role and the distinction from a paid care role, and how this influenced their perception and usage of terms such as “carer”.

> *“I think for a lot of people there is a kind of stigma around that word” (Participant 11)*

> *“I’m personally quite happy with the term but what annoys me is the fact that the press always use the term ‘carer’ to describe care workers…I think there needs to be a distinction between paid and unpaid people.” (Participant 6)*

Another difficulty for participants in identifying with the care-providing role was their experience of having care-related terminology and language used to describe them without this being explained, and before they had understood how their role had or might change. This led to some participants initially rejecting such terminology and this part of their identity. Some participants experienced an assumption or expectation from healthcare professionals that they would automatically adopt the caring role without consideration of their preferences.

> *“I mean they took it for granted that I was the wife of a husband who had a stroke.” (Participant 3)*

> *“There’s not an obligation to being a carer, but we feel obliged and feel that it is assumed to* be *within our role in the family to do that.” (Participant 5)*

> *“I think it was other people almost enforcing it on you, that you are now a carer, and you know, I didn’t feel like that.”(Participant 15)*

The lack of recognition or understanding from others, with regards to their changed role and identity, also comprised a barrier for people seeing themselves as a carer. Participants spoke about this lack of validation and acknowledgement leading to themselves struggling to acknowledge this new part of their identity.

> *“…a lot of family don’t really kind of register me* as *a carer, because they don’t really see a lot that goes on inside the normal day-to-day house.” (Participant 11)*

An important subtheme that directly informed the process of ‘how’ some participants related to the care role was *Cultural approaches to caregiving* (Subtheme 1.2.4). Some participants spoke about their cultural backgrounds shaping their understanding and approach to the caring role, and how it was perceived as a duty to their loved ones to provide this support. These participants identified as someone providing care and support, but this was in the context of being a friend or family member to the stroke survivor, and not a ‘carer’.

> *“All I can say is that the communities I come from, they see it as their duty and obligation and that’s it…that’s the kind of mentality we’ve all been brought up with.” (Participant 9)*

Furthermore, participants shared that different cultures and communities may not understand the concept of a ‘carer’, due to this not being a term used in their culture and therefore holding little meaning for them. Therefore, ‘how’ they came to self-identify as someone providing support or care was understood as a natural expectation within their culture. When drawing on their own culture and community, one participant described how English was not their first language, and that the term “carer” was not an established term or concept within their culture’s first language. They described how these terms are therefore difficult to relate to or engage with.

> *“…for people that do not have English as their first language, and they are caring for a friend or family member or elders, they wouldn’t even know what a ‘carer’ is.” (Participant 9)*

Exploring *when* participants started to identify with the care role is encompassed within the subtheme *Beginning to identify with the carer role* (Subtheme 1.2.3). The initial point of self-identification occurred at different points along the stroke trajectory for participants: an instant recognition at the time of the stroke event; realisation of the forthcoming changes in role and responsibility at point of planning discharge from hospital; on initial return home from hospital; and at a later stage when the stroke survivor had resided in the home environment, and they had been providing care for a while.

The timing of self-identification was influenced by the same factors as those pertaining to how participants reached the point of self-identification. Their exposure to these factors, and the timing of when they experienced them, presented as directly influencing when participants started to see themselves as providing care.

#### What care-related language have participants experienced and what are their preferences around this?

The second study aim was to explore participants’ preferences and experiences of care-related language. Relevant findings are mainly drawn from the theme of *Acceptability of care-related language and terminology* (Theme 4).

Participants presented two contrasting opinions regarding preferences and perceived acceptability of the term ‘carer’. One subgroup expressed a favourable view, feeling this term appropriately acknowledged their role in relation to their loved one, above that of what they perceived to be a typical relationship with that person. They perceived the term as broad, capturing the different elements the role may involve. This group also discussed a sense of duality in identifying with both a caring role and a relationship role with the stroke survivor, and the importance of acknowledging this additional role.

> *“…for me it felt like it was important for the distinction to be made between ‘carer’ and ‘partner’ and that both are equally as important…I say partner and carer because I feel like ‘carer’ was a bit like, well I’m not just her carer but I’m also her partner. But then I’m not just her partner, because I’m also her carer. So I kind of give myself a double title now.” (Participant 11)*

Another subgroup of participants held a contrasting view, rejecting care-related language in relation to describing themselves. Reasons for this are reflected in the previously discussed barriers for carer self-identification. Examples include participants feeling that using such terms may detract from or invalidate their relationship role with the person; holding a perception of the “carer” role that didn’t match their experiences; terminology not being culturally appropriate or accessible; and stroke survivors having a negative view of care-related language.

Regardless of individual preferences, participants spoke about feeling the need to use care-related language in a functional manner (Theme 5) to help others, such as healthcare professionals, understand their role and identity (Subtheme 5.1). They described using this language to promote inclusion and respect of their own opinions and viewpoints in the rehabilitation and support of the stroke survivor.

> *“I* would *use the terms carer and wife interchangeably, and which terms I use would depend on where I am and who I am talking to.”(Participant 3)*

> *“I would use [carer] generally to describe my role, yeah. If I needed to preface anything, you know like if I was speaking to a professional.”(Participant 5)*

Many participants discussed using terms such as ‘carer’ due to perceiving a lack of alternative, and feeling a sense of powerlessness, in that the term ‘carer’ is so widely used that they felt it would be challenging to change this or to make other people understand their own preferences if they differed from this (Subtheme 5.2).

> *“Like many other people I am sure, I have just accepted the word carer because that’s, you know, we’ve been* told *that’s what you are.” (Participant 9)*

#### What has the experience of support been for participants?

The final study aim was to explore participants’ experiences of support in their role of providing informal care. Theme 3 pertains to the participants’ perceptions and reflections on their own experiences of support. Participants discussed the types and sources of support that they had experienced (Subtheme 3.1). Multiple support sources were identified, including family and friends and local community such as neighbours, church and faith groups. A core source of support was connecting with other people who were also providing informal care after stroke. Participants who had not been able to access this peer support highlighted it as something they would have found beneficial.

> *“…just the classic thing of talking to people who are in the same situation as you and who understand, even if their particular circumstances are different they will get the general idea, and it just helps you realise that you’re not alone.” (Participant 11)*

Some participants discussed indirectly benefitting from the stroke survivor accessing support services, in terms of time for self-care and meaningful activities. Posters and informational resources were often helpful to participants when these were available. Participants described suggestions for improving future carer support provision (Subtheme 3.2). Participants felt improvements could be made in terms of signposting to resources and support, including peer support; accessibility of information, including printed, visual and alternative language formats of information; information on practical support (e.g. carer’s allowance); more variety in support options, including remote options for carers who are unable to attend in-person; improved communication between informal carers and healthcare services, including monitoring and reviewing their wellbeing and care needs throughout the stroke trajectory.

> *“I think that everybody needs to be made aware that this help is there for them. And that when they are ready, whether that is day one or not, and if they want to come back three months later, they can come back. We need to know it’s there for us when we are ready, because everybody’s ready differently I suppose, and you know, there will come a time where that person will realise they need that help now, and they need to know where they can find that help when they are ready and when they want it.” (Participant 11)*

All participants discussed their experiences around accessibility of support (Theme 2) and described factors that had facilitated their access to support for themselves (Subtheme 2.1). These factors included encouragement from other people to prioritise their own needs and access help; support services and information being readily available and in an accessible format; healthcare professionals asking about them and their wellbeing during contacts.

Participants also discussed factors that made it more difficult for them to access support (Subtheme 2.2). These included them struggling to recognise and prioritise their own needs; worrying about the consequences of them asking for help on the stroke survivor; having limited time to attend in-person support due to caring responsibilities; timing of support feeling inappropriate. Participants also acknowledged that factors such as geographical living location, council and service funding impacted their access to local resources and support.

> *“If you’re in a good relationship with your partner, it’s very hard to ask for help because I suppose in your mind, you’re frightened that they might be taken away from you.” (Participant 1)*

Language used around support was also raised as a barrier for participants accessing this.

> *“People don’t necessarily relate to carer. And most things are phrased as being for a carer, so it’s hard for people to accept resources if they don’t believe they fall under that category.” (Participant 13)*

## 4. Discussion

This study explored the self-identification process of informal carers, their experiences of care-related language and support. Through interviews with carers of stroke survivors, five main themes and related subthemes were identified that develop our understanding of how and when carers identify as such. Factors including the stroke severity and impact, and support from others to recognise their changed role, helped participants move towards identifying with their carer role. Barriers to self-identification included a perceived lack of time and space to process their own needs and changed role, and a lack of recognition from other people. One’s cultural background and beliefs were identified as shaping someone’s perception and interpretation of the care role. Care-related language usage interestingly presented as intersecting with the process of carer-self-identification, as well as the perceived accessibility of support provision. Findings will be discussed within the context of the wider relevant literature, and associated clinical implications summarised.

Study findings indicate the timing of self-identification differs between individuals, yet there are core time points along the stroke trajectory when self-identification can become apparent: at the time of the stroke event, at the point of discharge planning, on initial return home, and after a period of longer-term caring. Previously established theories map stroke informal carers’ changing needs and experiences at core points along the stroke trajectory [17]. These core points are largely reflective of those identified by participants in the present study as times of self-identification.

Results from the present study illustrate the varying depictions held by participants around what a “carer” is, which in turn influenced how this formed, or did not form, part of their identity. Identity theory positions the roles that people occupy as the basis of identity, whilst social identity theory places emphasis on social structures, such as group belonging, as a source of identity [41]. It can also be the case that multiple identities can be occupied simultaneously, with different identities either reinforcing or conflicting with each other [42]. This was apparent in the present study, with some participants feeling more aligned with either the relationship or caring role at different points. It was also evident that most participants identified as simultaneously occupying a relationship role (e.g. son, granddaughter, spouse) and ‘carer’ role. Participants adopted the identity of ‘carer’ to varying extents, which is consistent with other informal caregiving research in other populations indicating relatives’ affiliation with the term ‘carer’ being variable [43].

Participants who did not identify as a ‘carer’ commonly expressed having a preconceived idea of what a ‘carer’ was, which did not correlate with their experiences of informal caregiving, leading to them struggling to integrate or accept the carer part of their identity. Participants also often conflated an ‘informal carer’ with a paid or employed carer, which contributed to this. Additionally, some participants described the term ‘carer’ being used to describe them without explanation or discussion, which caused increased confusion and misalignment with their identity, particularly for individuals who were new to providing informal care. This highlights the need for provision of educational support to aid the understanding of what a ‘carer’ is, who this term refers to when used by healthcare professionals and emphasising how the term ‘carer’ can be used to reflect numerous individuals and situations [3]. This is a clinical recommendation which could be addressed by healthcare professionals and third sector organisations aimed at supporting informal carers. The wider family could also be supported to understand the primary informal carer’s role, as this is evidenced to enhance the provision of positive family support for the informal carer [44].

Being involved in the rehabilitation processes, and thus understanding the extent of support needed by their loved one, was identified by participants as a facilitator to moving towards a position of self-identification and acknowledgement that they may need to take on a care-related role. Participants who had not experienced this often identified with the care role later, such as on discharge, and described this as a more sudden transition to the role. This aligns with existing literature highlighting the need for services to support carers with this role transition, including early preparation and involvement in the rehabilitation processes [45]. Therefore, it is recommended that services involve carers and family members of stroke survivors in rehabilitation processes.

Participants spoke about other individuals inappropriately using care-related language to describe them and seemingly making assumptions that participants would adopt the carer role, which sometimes led to them rejecting this part of their identity. A distinction was described between ‘carer’ as an incorporated component of self-identity versus ‘carer’ being used as a label to describe or refer to them. An intersection was apparent between the two in the present study. Participants who had reached the point of identifying with their care role or ‘carer identity’, presented as more comfortable with this part of their identity being acknowledged through care-related language. Some discussed benefitting from this identity being acknowledged. By contrast, participants who did not affiliate with the care-providing identity found care-related language less acceptable and appropriate to be used to describe them. The incongruous use of care-related language with one’s sense of identity also likely contributed to a sense of disempowerment for carers, as they outlined how they did not feel able to express their own terminology preferences. The Caregiver Readiness Framework outlines the importance of assessing individuals’ capacity and willingness to undertake a care role early in the stroke trajectory, and not making assumptions about their role in the stroke survivor’s care [45]. Informal caregivers can at times experience reduced confidence and self-esteem in themselves and their role [46–47], and individuals who have expectations placed on them that they *should* undertake the care role often reject this and feel uncomfortable [13]. Therefore, the corresponding clinical recommendation is that assumptions should not be made about if or to what extent someone will take on the care role.

As previously mentioned, literature exploring the effectiveness of support for informal caregivers of stroke survivors highlights that these individuals often do not see themselves as ‘carers’, which can contribute to difficulties with them engaging with staff for carer-focussed support [11]. This finding was reinforced and further explored in the present study. Participants shared that they felt if someone did not identify as a ‘carer’ that they would be less inclined to engage with support using care-related terms, as they may feel the support was not aimed at them. An implication for clinical practice is that more accessible and inclusive language should be used when designing and implementing support programmes, as this may improve engagement from those not identifying as ‘carers’.

The findings of the study highlighted how current support provision is not always accessible to individuals from different communities and cultures. Examples included using terms such as ‘carer’ that do not resonate with these groups and having information in inaccessible formats. Furthermore, it is well evidenced in the literature that ethnic minority groups, such as people of Black or South Asian ethnicities, are disproportionately affected by stroke [48–49]. Synthesising these two findings, it is important to ensure support services are accessible to and reflect the needs and values of people from a variety of cultural and ethnic backgrounds. The present study also indicated that the format of support doesn’t often correlate to the needs of the carer, such as care responsibilities making in-person support inaccessible. Other participants suggested information should be presented in more varied formats, including printed or visual instead of solely verbal. In summary, it is recommended that the design and implementation of support considers the accessibility in terms of potential language and cultural factors, and different sensory and information processing needs of carers.

In exploring *when* informal carers come to identify with the care role, study findings showed that there are different time points at which individuals reach this, which can inform the timing of when support is offered. As participants identify at different points along the stroke trajectory, it is important that they are aware of what support is available to them when they need or may benefit from this. The facilitators identified through the study of helping individuals reach a point of identifying with the care role should be considered and implemented where possible. This includes supporting individuals to connect with other informal carers, and to recognise and prioritise their own needs. Established frameworks around caregiving in stroke show that carers have support needs from as early as the stroke event [17, 50], and so support should be offered from this point and signposting to community and longer-term support options as needed.

Finally, findings emphasise the importance of maintaining an active awareness of informal carers, their experiences and needs, both at a service level and more widely in research. At an individual level, participants believed they would feel more supported if there was improved communication between themselves and the healthcare professionals, including monitoring and checking in with their wellbeing and sense of coping. It is well established that informal carers often struggle to identify and prioritise their own needs [18, 51] so this clinical recommendation of reviewing the wellbeing of carers should address this. Furthermore, the existing literature continues to highlight a need for further research into the experience of informal carers as there remains much to be explored. In line with existing literature, this study reinforces the continued need for carer-focussed research within the stroke population.

### 4.1. Strengths, limitations and future research

To the research team’s knowledge, this is the first study to explore the self-identification process of stroke informal carers. Study strengths include the qualitative approach, which allowed elicitation of the richness and complexities of carers’ lived experiences through interviews [52]. The sample heterogeneity in terms of age and relationship to the stroke survivor, and duration of care experience also strengthened the study. Collaboration with a formed PCPI group promoted the study accessibility and appropriateness to participants. However, the study is limited by its transferability of findings to different cultures and ethnic populations, due to the limited diversity in participant ethnicity. A more diverse sample should be obtained to explore this specific area in future research. Alternatively, this topic area could be explored within specific cultures that are often underrepresented in research but disproportionately affected by stroke. Furthermore, the template analysis approach effectively identified factors relevant to if and how someone sees themselves as a carer. The incorporation of deductive and inductive approaches was helpful for answering the research questions: the deductive component aided the inductive data analysis, through acknowledging existing knowledge to support data interpretation. Future research could adopt interpretative phenomenological analysis approaches, to provide insights into what the experience of being a carer is like, and how one makes sense of this identity. The retrospective approach to the areas explored in this study allowed participants to reflect on their caregiving experience and perspectives they developed along the way. However, studying the process of self-identification and adjustment to the carer role prospectively may allow the critical and gradual processes of change and transition to be explored as they are unfolding [53].

### 4.2. Conclusions

This novel study offers an understanding of how and when informal carers of stroke survivors identify with this role and the factors influencing this process. The experience of care-related language and the extent to which it comprises a barrier, alongside other factors, for carers accessing support is also established. This study provides key clinical implications to potentially improve the future support provision for this population.

## Data Availability

All data produced in the present work are contained in the manuscript

## 5. Declarations

### Ethics approval and consent to participate

Ethics approval for this study was received by the University of Manchester Research Ethics Committee. Informed consent was gained from all participants prior to their participation in the study.

### Consent for publication

All participants gave informed consent for their anonymised interview data to be used in publications.

### Availability of data and materials

All participants provided informed consent for their anonymised interview transcript being uploaded to the University of Manchester’s depository and made available to other researchers. This data is not publicly available due to potential compromising of individuals’ privacy, and explicit consent for this not being sought.

### Author contributions

All authors conceptualised and participated in the design of this study. BH led on coordination of recruitment, with the support of the other authors (AB and RB). BH coordinated the data collection and performed the template analysis of data. All authors were involved in and reflected on the interpretation of findings and conclusions drawn.

BH prepared the first and subsequent manuscripts. AB and RB provided invaluable academic supervision throughout the project, provided guidance on the data analysis process and revisions of drafts, and contributed to the synthesis and interpretation of the data.

## Acknowledgements

The authors wish to express their thanks and gratitude to all of the individuals who took part in our study, and shared their time and experiences. Special thanks goes to the members of the carer advisory group who provided invaluable support and perspectives to the research team throughout the study. Thank you all, for making this study possible. We would also like to thank all the organisations and individuals that supported with recruitment to this study, and allowed us to share our findings with them.

