## Supplemental FIle 1 for "Informal caregiving following stroke: a qualitative exploration of carer self-identification, care-related language and support experiences"

**Final Coding Template**

1. Adopting and adjusting to the care role

1.1. Adjusting to the dynamic role of a carer

1.2. Identifying with the carer role

1.2.1. Facilitators to self-identification

1.2.2. Barriers to self-identification

1.2.3. Beginning to identify with the carer role

1.2.4. Cultural approaches to caregiving

1.3. Learning new skills and responsibilities

1.4. Advocacy and driving change

1.5. Positive aspects of caregiving

2. Accessibility of support

2.1. Facilitators to accessing support

2.2. Barriers to accessing support

3. Perceptions of support

3.1. Types and sources of support

3.2. Suggestions for improving support provision

4. Acceptability of care-related language and terminology

5. Function of care-related language and terminology

5.1. Increasing others’ understanding of own identity

5.2. Sense of powerlessness to change care-related language use
