## Supplemental File 2 for "Informal caregiving following stroke: a qualitative exploration of carer self-identification, care-related language and support experiences"

**A priori theme development and refinement process**

A priori theme names were either modified to be more reflective of the interview data, or did not feature within the final coding template and were instead captured within a broader theme or subtheme

**Initial tentative a priori themes**

(derived from reviews of themes and concepts prevalent in relevant existing literature)

Expectations of caregiving

Uncertainty and unpredictability

Increased responsibility

Changing support needs

Communication

Role change

Theme 1: Adopting and adjusting to the care role

Subtheme 3.2: Suggestions for improving support provision

Subtheme 1.3: Learning new skills and responsibilities

Subtheme 1.1: Adjusting to the dynamic role of a carer

Subtheme 1.2.1: Facilitators to self-identification

Subtheme 1.2.2: Barriers to self-identification

**Themes and subthemes of final coding template**

Theme development
